# Changes in Aortic Centerline Length and Curvature Predict Aortic Diameter Growth in Chronic Type B Aortic Dissection

**DOI:** 10.1101/2025.10.15.25334577

**Authors:** Xue Liang, Hai Dong, Marc-Philipp H. Schmid, Minliang Liu, Hannah L. Cebull, Michael Zhang, Sunny Xu, Muhammad Naeem, John N. Oshinski, John A. Elefteriades, Rudolph L. Gleason, Bradley G. Leshnower

## Abstract

**Objective:** Aortic diameter is the primary metric used to trigger surgical intervention in chronic Type B aortic dissection (TBAD) This study investigates the correlation between diameter growth and changes in aortic centerline length, curvature and aortic volume in chronic TBAD.

**Methods:** We retrospectively collected 80 computed tomography (CT) images from 31 chronic TBAD patients. Eighteen patients had three follow-up CT images (CT-1, CT-2, and CT-3), while the remaining 13 had two scans (CT-1 and CT-2). Three-dimensional geometry and aortic centerlines were reconstructed from each CT scan, based on which the mean and maximum TBAD diameter were obtained. The centerline length (CL) and curvature (CC) from the left subclavian to celiac branch were also obtained. The relationship between the diameter growth rate vs. CL/CC growth rate were investigated using linear regression analysis.

**Results:** Between CT-1 & CT-2, the CL growth rate had an inverse correlation with the mean (*R* = −0.7923, *p* < 0.001) and maximum (*R* = −0.7880, *p* < 0.001) diameter growth rate. The CC growth rate also had an inverse correlation with the mean (*R* = −0.8304, *p* < 0.001) and maximum (*R* = −0.8430, *p* < 0.001) diameter growth rate. Between CT-2 & CT-3, we found similar inverse correlations between diameter growth rate vs. CL/CC growth rate (all |*R*| > 0.78, *p* < 0.001). Moreover, early CL/CC growth (CT-1 & CT-2) predicted later diameter growth (CT-2 & CT-3) more strongly (|*R*| > 0.72) than did early diameter growth itself (|*R*| < 0.61), indicating the centerline length/curvature may be better predictors for diameter growth of chronic TBAD.

**Conclusions:** In chronic TBAD, the CL/CC growth has an inverse relationship with aortic diameter growth, which may be used to predict diameter of chronic TBAD.

**Central Picture:** 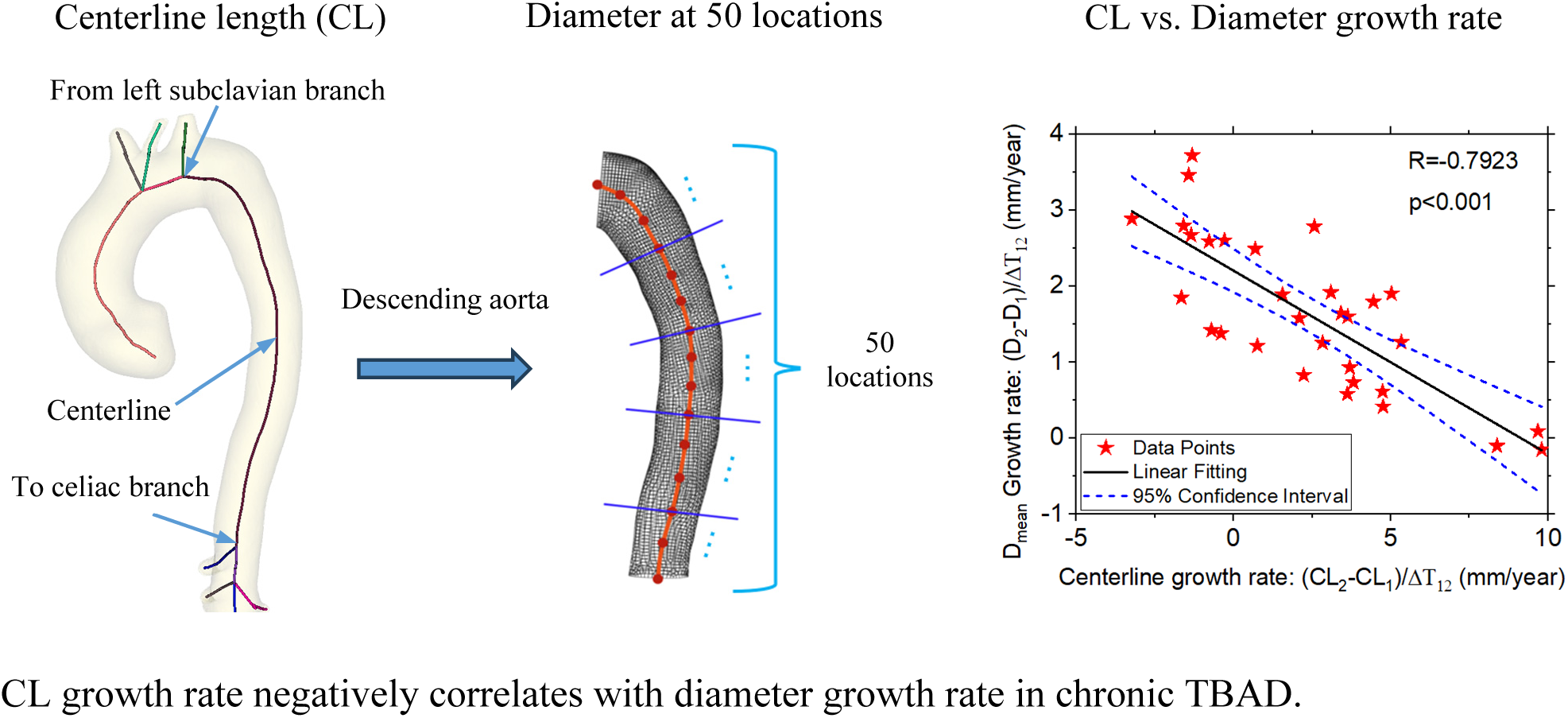

**Central Message:** In chronic Type B Aortic Dissection (TBAD), the growth of centerline length and curvature has an inverse correlation with the diameter growth, which may be used to predict chronic TBAD diameter.

**Perspective Statement:** This study investigates the correlation between diameter growth and changes in aortic centerline length (CL) and curvature (CC) in chronic TBAD. Based upon our findings, the CL/CC growth has an inverse correlation with the diameter growth, and early CL/CC growth predicted later diameter growth more strongly than did early diameter growth itself.

## 1. INTRODUCTION

Optimal medical therapy (OMT) with surveillance imaging remains the primary therapy for patients with uncomplicated TBAD^1–4^. This strategy produces excellent short-term survival, but poor long-term outcomes with 5-year intervention-free survival rates < 50%^5^. Prophylactic TEVAR has been proposed as an alternative in these patients^6^, but carries the risk of stroke, paraplegia, retrograde type A aortic dissection^7,8^ and results in aortic stiffening that can result in adverse effects on the blood pressure and cardiac function^9–11^. Several studies have searched for anatomic predictors of growth in TBAD based upon analysis of cross-sectional imaging, but aortic diameter remains the only consistent metric that has been validated across multiple studies^12–14^.

Ideally, uncomplicated TBAD patients who will subsequently develop large false lumen aneurysms could be identified early in the acute or subacute phases when the mechanical properties of the dissection flap remain favorable for aortic remodeling and undergo TEVAR. Unfortunately, attempts to define clear anatomic predictors of OMT failure have been largely unsuccessful, and aortic diameter remains the lone predictor that has been consistently identified and validated across multiple studies^12,15,16^.

The inability to identify conventional anatomic predictors of OMT failure has sparked investigations into aortic morphology, with the hypothesis that novel characteristics such as aortic length and curvature could predict adverse aortic events (rupture, dissection and death)^17–21^. In a cohort of 522 ascending thoracic aortic aneurysms patients, the Yale group demonstrated that ascending aortic length was strongly associated with adverse aortic events^17^. Sun and colleagues reported that the combination of aortic length and curvature may serve as a sensitive predictive tool for the development of acute Type A aortic dissection in patients with ascending aortic aneurysm diameters < 5.0 cm^22^. Additional data from the same group demonstrated that aortic angulation, arch type and tortuosity were associated with the development of acute TBAD. These findings highlight the relevance of investigating aortic morphology in TBAD as a risk factor for FL aneurysm expansion and adverse aortic events.

Currently there is a paucity of data examining the impact of aortic morphology on aortic diameter growth rates for patients with uncomplicated chronic TBAD treated with OMT. The purpose of this study is to investigate the relationship between diameter growth rate and changes in aortic centerline length and curvature in chronic TBAD patients. By analyzing these relationships, we aim to improve current risk assessment methods, that may enable early identification of patients at high risk for FL aneurysm expansion and adverse aortic events.

## 2. DATA AND METHODS

This study was approved by the Emory University Institutional Review Board (IRB00109646 and STUDY00002737). Patients’ informed written consents were obtained for the publication of the study data. A retrospective review of the Emory aortic database was conducted for patients who presented with acute uncomplicated TBAD and were treated with OMT between 2007 and 2025. Inclusion criteria for this study consisted of patients in the chronic phase of TBAD with their initial diagnostic computed tomography (CT) scan (CT-1) with contrast and at least one follow-up scan available for analysis. This review yielded 31 patients with a total of 80 CT scans. Eighteen (58%) patients had 2 follow-up scans (CT-2, CT-3), and the remaining 13 patients had a single follow-up scan (CT-2) available for analysis.

### 2.1 Reconstruction of TBAD geometry and centerline

Three-dimensional (3D) geometry (Fig. 1a, b) of the dissected aorta including both true and false lumens was reconstructed from each of the 80 CT scans using 3D Slicer (www.slicer.org). Next, a line was semi-automatically created within the 3D aortic reconstruction that represented the medial axis of the aorta, known as the aortic centerline (Fig 1a). The purpose of the aortic centerline was to facilitate accurate aortic diameter measurements and evaluate aortic growth in the longitudinal direction. It is well recognized that the most accurate method of aortic diameter measurement is with a cross-sectional plane that is taken perpendicular to the medial axis of the aorta^23^. Using a multiplanar reconstruction the aorta can be unfolded into a straight tube along its centerline to allow for accurate measurement of aortic growth in the longitudinal direction via centerline length.

**Fig. 1.**
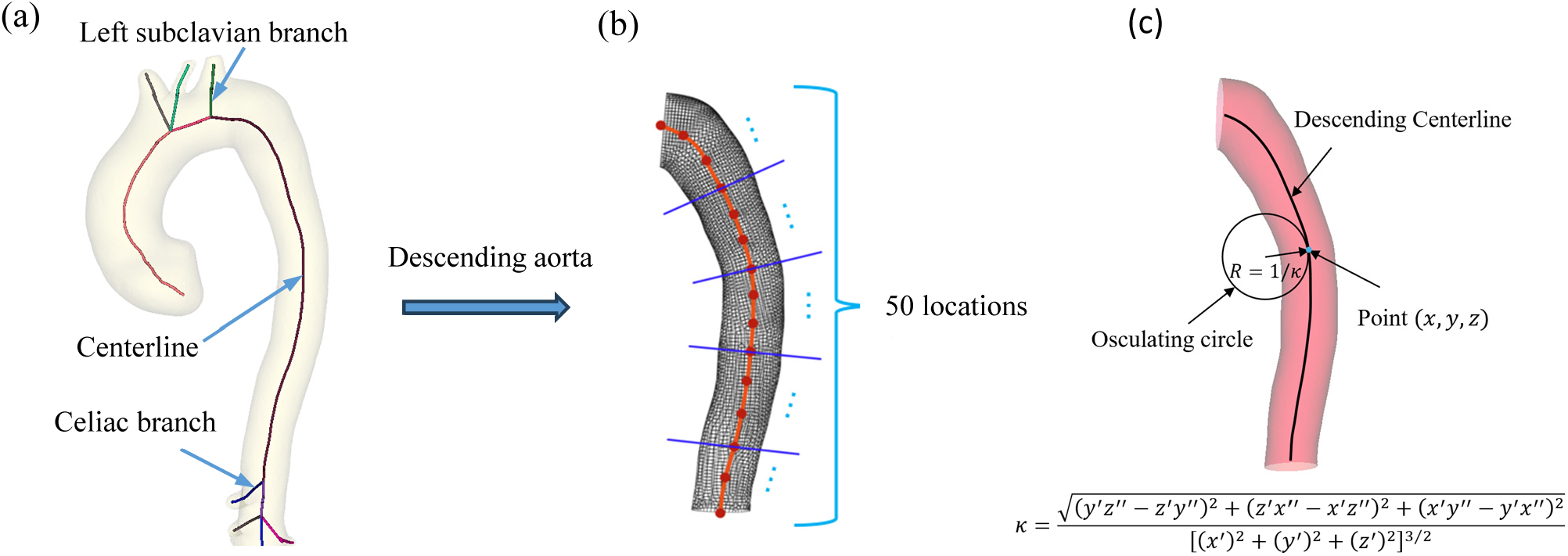
(a) Three-dimensional geometry and centerline of the aortic outer wall constructed from CT images; (b) Outer wall of descending aorta: Diameter at 50 locations, evenly distributed along the centerline, were obtained; (c) Calculation of the local curvature (*κ*) of a point with coordinates (*x, y, z*). The radius (*R*) of the osculating circle is the reciprocal of *K*.

### 2.2 Calculation of annual diameter growth rate

For each 3D aortic reconstruction, the dissected descending thoracic aortic segment (left subclavian artery to celiac artery) was extracted (Fig. 1b) for aortic diameter and centerline analysis. The diameters at 50 locations along the descending thoracic aorta, evenly distributed along the centerline (Fig. 1b), were obtained for each of the 80 CT scans using customized MATLAB codes. Based upon these 50 measurements for each CT scan, the mean and maximum diameters of the descending thoracic aorta were calculated. The annual diameter growth rate (mean and maximum) between {CT-1 & CT-2}i and {CT-2 & CT-3} were calculated by dividing the diameter growth with the time interval (in years) between each of the two scans.

### 2.3 Calculation of centerline length and curvature growth

Based on the aortic centerline constructed in Section 2.2 (Fig. 1a), the centerline length (CL) between the left subclavian artery and the celiac artery was measured using as straightened multiplanar aortic reconstruction for all 80 CT scans. Additionally, a curved multiplanar aortic reconstruction was used to calculate the mean and maximum centerline curvature (CC, inverse of local radius, Fig. 1c). Specifically, a centerline can be characterized by the function **r**(*t*) = {*x*(*t*), *y*(*t*), *z* (*t*)}, where *t* is the arc length (0 ≤ *t* ≤ *L*) and **r**(*t*) is one point on the centerline with the coordinate of {*x*(*t*), *y*(*t*), *z* (*t*)}. Then, the local curvature *κ*(*t*) at the point **r**(*t*) is defined _as_^24^

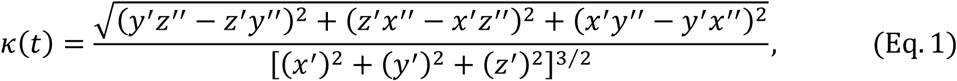

where (·)′ and (·)′′ are the first and second derivatives with respect to the arc length *t*. To ensure accuracy, each centerline was discretized with a minimum number of 115 points. The coordinates of each point (*x*_i_, *y*_i_, *z*_i_), *i*=1,2,..115, are exported from the 3D Slicer (www.slicer.org). The corresponding 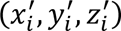 and 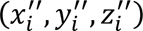 are subsequently calculated using the *gradient* function in MATLAB2024a (Mathworks Inc., Natick, MA). The local curvature *K*_i_ of each point (*x*_i_, *y*_i_, *z*_i_) was obtained by substituting 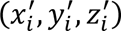 and 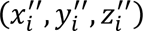 into (Eq. 1). The annual CL growth rates between {CT-1 & CT-2} and between {CT-2 & CT-3} were calculated by dividing the CL growth by the time interval (in years) between each of the two scans. The mean and maximum annual growth rates of CC were calculated using the same method.

### 2.4 Calculation of aortic volume

For each CT scan, the descending thoracic aortic volume was quantified from the 3D segmentation using the 3D Slicer software to preserve only the descending aorta from the left subclavian artery to the celiac artery. The enclosed volume was obtained by summing the volumes of all voxels contained in the region, using ‘Scalar Volume’ in the module of ‘Segment Statistics’ in 3D Slicer (www.slicer.org) for all 80 CT scans.

### 2.5 Linear regression and statistical analysis

Linear regression was applied to investigate the correlation between the diameter growth and the CL and CC growth. The strength of the linear correlation was quantified by the Pearson correlation coefficient (*R*), and statistical significance was set at *p* < 0.05, with the values of *R* and *p* calculated with the *corrcoef* function in MATLAB 2024a (Mathworks Inc., Natick, MA).

## 3. RESULTS

### 3.1 Patient data

Patient demographics of the 31 patients involved in this study are provided in Table 1. The cohort had a mean age of 56.84 years and included 19 male (61.29%). Hypertension was present in all patients (31/31, 100%). Additional comorbidities included congestive heart failure in 5 (16.13%) patients, prior myocardial infarction in 1 (3.23%) patient, and prior cerebrovascular accident in 1 (3.23%) patient. Two patients (6.45%) had a connective tissue disorder, 2 patients (6.45%) reported a family history of aortic dissection, and 2 (6.45%) patients reported cocaine use. The median interval between surveillance scans was 23.00 months between CT-1 and CT-2, 19.75 months between CT-2 and CT-3, and 42.50 months between CT-1 and CT-3.

**Table 1.** Demographics of the patients involved in this study

|  | All Patients<br>N=31 |
| --- | --- |
| Age (years) – mean (SD) | 56.84 (±11.20) |
| Male sex – no. (%) | 19 (61.29%) |
| Connective tissue disease – no. (%) | 2 (6.45%) |
| Hypertension – no. (%) | 31 (100%) |
| Congestive heart failure – no. (%) | 5 (16.13%) |
| Myocardial infarction – no. (%) | 1 (3.23%) |
| Cerebrovascular accident – no. (%) | 1 (3.23%) |
| Family history – no. (%) | 2 (6.45%) |
| Cocaine use – no. (%) | 2 (6.45%) |
| Max descending diameter (cm) – median [IQR] | 5.52-4.02[1.22] |
| Median time between CT-1 and CT-2 (month) | 23.00 |
| Median time between CT-2 and CT-3 (month) | 19.75 |
| Median time between CT-1 and CT-3 (month) | 42.50 |

### 3.2 Centerline length growth vs. diameter growth

The absolute values of the aortic diameter, centerline length (CL) and curvature (CC) for all 80 scans were reported in Table 2. The CL growth rate was found to have an inverse correlation with both the mean (*R* = −0.7923, *p* < 0.001) and maximum (*R* = −0.7880, *p* < 0.001) diameter growth rates between CT-1 and CT-2 (Fig. 2a, b). Similar correlations between CL growth rate vs. mean (*R* = −0.7844, *p* < 0.001) and maximum (*R* = −0.8146, *p* < 0.001) diameter growth rate were found in the time period between CT-2 and CT-3 (Fig. 2c, d). The CL growth rate calculated between CT-1 and CT-2 also has an inverse correlation with the mean (*R* = −0.7825, *p* < 0.001) and maximum (*R* = −0.7203, *p* < 0.001) diameter growth rate between CT-2 and CT-3 (Fig. 2e, f), indicating that the CL growth (i.e. aortic elongation) may be used to predict aortic diameter growth.

**Fig. 2.**
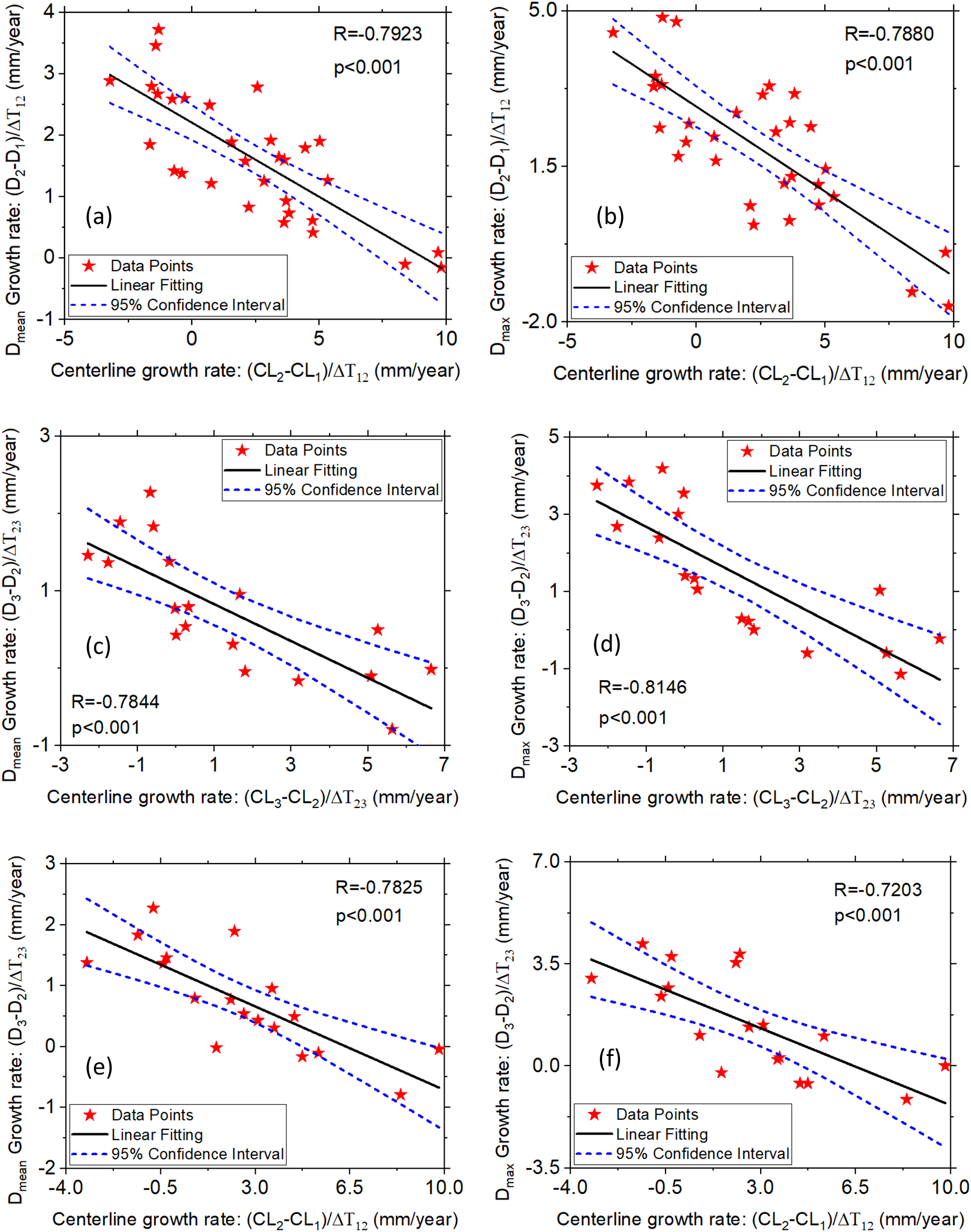
(a, b): Correlations between the centerline length (CL) growth rate and mean/maximum (a/b) diameter (D) growth rate during Scan-1&Scan-2; (c, d) Correlations between the CL growth rate and mean/maximum (c/d) diameter growth rate during Scan-2&Scan-3; (e, f) Correlations between the CL growth rate (Scan-1&Scan-2) and mean/maximum (e/f) diameter growth rate (Scan-2&Scan-3).

**Table 2.** Data of Aortic Diameters, Centerline Lengths, and Centerline Curvature in 31 Uncomplicated TBAD patients. Delta represents the increment from the first CT to the last CT.

| Patient ID | Max Diameter (mm) |  |  |  | Centerline Length (mm) |  |  |  | Max Centerline Curvature (mm<sup>-1</sup>) |  |  |  |
| --- | --- | --- | --- | --- | --- | --- | --- | --- | --- | --- | --- | --- |
|  | CT-1 | CT-2 | CT-3 | Delta | CT-1 | CT-2 | CT-3 | Delta | CT-1 | CT-2 | CT-3 | Delta |
| 1 | 31.28 | 31.96 | 32.48 | 1.20 | 234.76 | 239.20 | 241.75 | 6.99 | 0.15 | 0.18 | 0.18 | -0.10 |
| 2 | 40.17 | 46.81 | 55.05 | 14.88 | 290.78 | 299.82 | 299.81 | 9.03 | 0.21 | 0.14 | 0.11 | 0.23 |
| 3 | 36.80 | 42.88 | 41.69 | 4.89 | 268.60 | 279.90 | 290.41 | 21.81 | 0.23 | 0.16 | 0.46 | 0.35 |
| 4 | 29.14 | 28.53 | 27.39 | -1.75 | 231.65 | 235.50 | 241.12 | 9.47 | 0.21 | 0.32 | 0.56 | 0.19 |
| 5 | 37.39 | 40.10 | 41.25 | 3.86 | 300.25 | 301.51 | 301.85 | 1.60 | 0.15 | 0.15 | 0.34 | 0.72 |
| 6 | 43.93 | 41.45 | 41.48 | -2.45 | 246.06 | 260.76 | 266.53 | 20.47 | 0.12 | 0.38 | 0.84 | 0.48 |
| 7 | 32.37 | 34.46 | 33.49 | 1.12 | 274.50 | 283.58 | 288.76 | 14.26 | 0.08 | 0.13 | 0.56 | -0.52 |
| 8 | 42.68 | 43.16 | 46.85 | 4.17 | 260.71 | 266.47 | 265.07 | 4.36 | 0.61 | 0.20 | 0.09 | -0.44 |
| 9 | 34.90 | 37.91 | 44.94 | 10.04 | 253.91 | 251.75 | 251.34 | -2.57 | 0.79 | 0.57 | 0.35 | -0.38 |
| 10 | 32.87 | 36.72 | 33.39 | 0.52 | 270.57 | 269.82 | 267.97 | -2.60 | 0.65 | 0.37 | 0.27 | -0.02 |
| 11 | 27.46 | 34.68 | 34.57 | 7.11 | 249.50 | 253.65 | 256.97 | 7.47 | 0.40 | 0.33 | 0.38 | -0.18 |
| 12 | 49.02 | 50.25 | 53.22 | 4.20 | 280.93 | 280.79 | 278.97 | -1.96 | 0.50 | 0.39 | 0.32 | -0.28 |
| 13 | 50.27 | 51.29 | 53.96 | 3.69 | 273.97 | 277.46 | 277.43 | 3.46 | 0.47 | 0.20 | 0.19 | 0.31 |
| 14 | 48.68 | 49.27 | 50.20 | 1.52 | 291.51 | 299.05 | 305.67 | 14.16 | 0.84 | 0.91 | 1.15 | -0.83 |
| 15 | 38.68 | 49.32 | 51.55 | 12.87 | 274.11 | 282.90 | 283.31 | 9.20 | 0.99 | 0.24 | 0.16 | 0.75 |
| 16 | 40.33 | 48.87 | 49.83 | 9.50 | 288.67 | 313.48 | 318.29 | 29.62 | 1.17 | 1.45 | 1.92 | -0.88 |
| 17 | 38.34 | 49.21 | 58.63 | 20.29 | 273.45 | 269.06 | 267.73 | -5.72 | 1.22 | 0.45 | 0.34 | -0.61 |
| 18 | 36.54 | 39.31 | 46.39 | 9.85 | 233.97 | 233.51 | 231.51 | -2.46 | 0.73 | 0.59 | 0.12 | -1.00 |
| 19 | 32.96 | 47.53 | ---- | 14.57 | 225.98 | 218.67 | ---- | -7.31 | 1.18 | 0.18 | ---- | 0.02 |
| 20 | 47.10 | 49.20 | ---- | 2.10 | 273.55 | 280.87 | ---- | 7.32 | 0.54 | 0.56 | ---- | -0.39 |
| 21 | 39.71 | 47.29 | ---- | 7.58 | 291.80 | 298.29 | ---- | 6.49 | 0.55 | 0.16 | ---- | -0.04 |
| 22 | 50.33 | 51.32 | ---- | 0.99 | 278.92 | 278.32 | ---- | -0.60 | 0.29 | 0.25 | ---- | -0.72 |
| 23 | 30.68 | 41.20 | ---- | 10.52 | 313.79 | 310.94 | ---- | -2.85 | 0.82 | 0.10 | ---- | 0.01 |
| 24 | 44.19 | 56.71 | ---- | 12.52 | 290.74 | 285.63 | ---- | -5.11 | 0.41 | 0.42 | ---- | -0.24 |
| 25 | 51.39 | 54.73 | ---- | 3.34 | 329.77 | 330.84 | ---- | 1.07 | 0.38 | 0.14 | ---- | -0.20 |
| 26 | 38.24 | 41.38 | ---- | 3.14 | 275.47 | 279.29 | ---- | 3.82 | 0.60 | 0.40 | ---- | -0.04 |
| 27 | 40.32 | 52.03 | ---- | 11.71 | 279.99 | 297.08 | ---- | 17.09 | 0.18 | 0.14 | ---- | 0.42 |
| 28 | 50.46 | 49.62 | ---- | -0.84 | 298.59 | 317.14 | ---- | 18.55 | 0.12 | 0.54 | ---- | 0.45 |
| 29 | 55.23 | 59.41 | ---- | 4.18 | 294.25 | 325.96 | ---- | 31.71 | 0.17 | 0.62 | ---- | -0.18 |
| 30 | 46.58 | 48.64 | ---- | 2.06 | 296.12 | 295.19 | ---- | -0.93 | 0.78 | 0.60 | ---- | -0.05 |
| 31 | 45.89 | 47.00 | ---- | 1.11 | 269.94 | 273.35 | ---- | 3.41 | 0.36 | 0.31 | ---- | 0.03 |

### 3.3 Centerline curvature growth vs. diameter growth

For the time period between CT-1 and CT-2, the mean and maximum centerline curvature (CC) growth rates had inverse correlations with the mean (*R* = −0.8304, *p* < 0.001) and maximum (*R* = −0.8430, *p* < 0.001) diameter growth rates, respectively (Fig. 3a, b). For the period between CT-2 and CT-3, inverse correlations between the mean/maximum CC growth rate vs. the mean/maximum (*R* = −0.8383/−0.8454, both *p* < 0.001) diameter growth rates were observed (Fig. 3c, d). Moreover, the mean and maximum CC growth rates in CT-1 and CT-2 also had inverse correlations with the mean (*R* = −0.8151, *p* < 0.001) and maximum (*R* = −0.8180, *p* < 0.001) diameter growth rate between CT-2 and CT-3 (Fig. 3e, f).

**Fig. 3.**
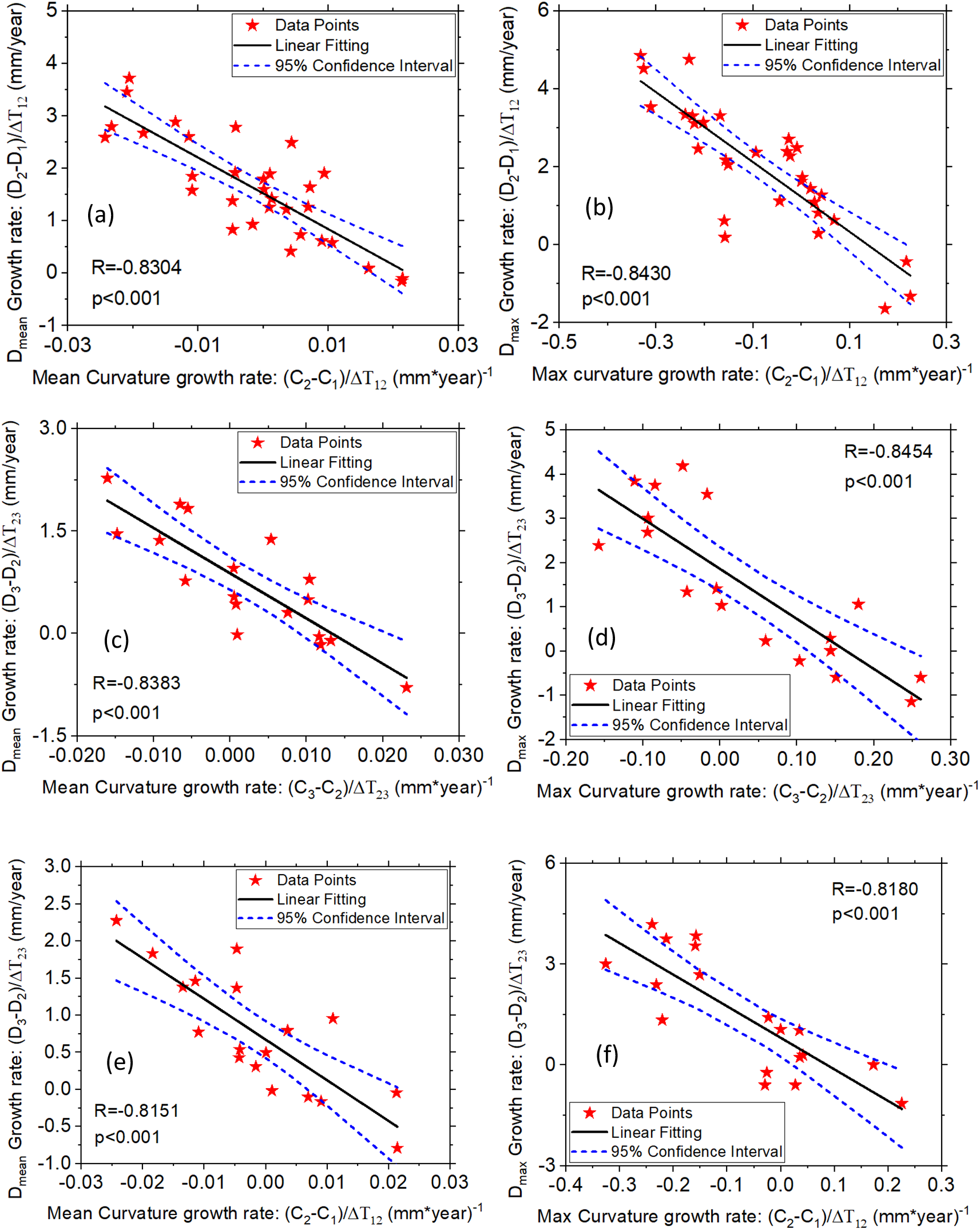
(a, b): Correlations between the mean/maximum centerline curvature (CC) growth rate and mean/maximum (a/b) diameter (D) growth rate during Scan-1&Scan-2; (c, d) Correlations between the mean/maximum (c/d) CC growth rate and mean/maximum diameter growth rate during Scan-2&Scan-3; (e, f) Correlations between the mean/maximum (e/f) CC growth rate (Scan-1&Scan-2) and mean/maximum diameter growth rate (Scan-2&Scan-3).

By comparing the results in Fig. 2 and in Fig. 3, the CC growth rate has a stronger inverse correlation with diameter growth than that of the CL growth rate, which suggested that the CC growth may be a better predictor for the diameter growth than the CL growth.

### 3.4 Centerline length (CL) growth vs. centerline curvature (CC) growth

We also investigated the relationship between the CL growth rate and the CC growth rate. We found that, for both time periods between CT-1 & CT-2 and CT-2 & CT-3, the CL and CC growth rates have positive correlations (Fig. 4 a-d). For the period of CT-1 & CT-2 (Fig. 4a, b), the correlation coefficient (*R*) was 0.8559 and 0.8382 for CL growth rate vs. mean and maximum CC growth rates, respectively (p< 0.001). For the period of CT-2 & CT-3 (Fig. 4c, d), the correlation coefficient (*R*) was 0.7256 and 0.6927 (p<0.01) for CL growth rate vs. mean and maximum CC growth rates, respectively.

**Fig. 4.**
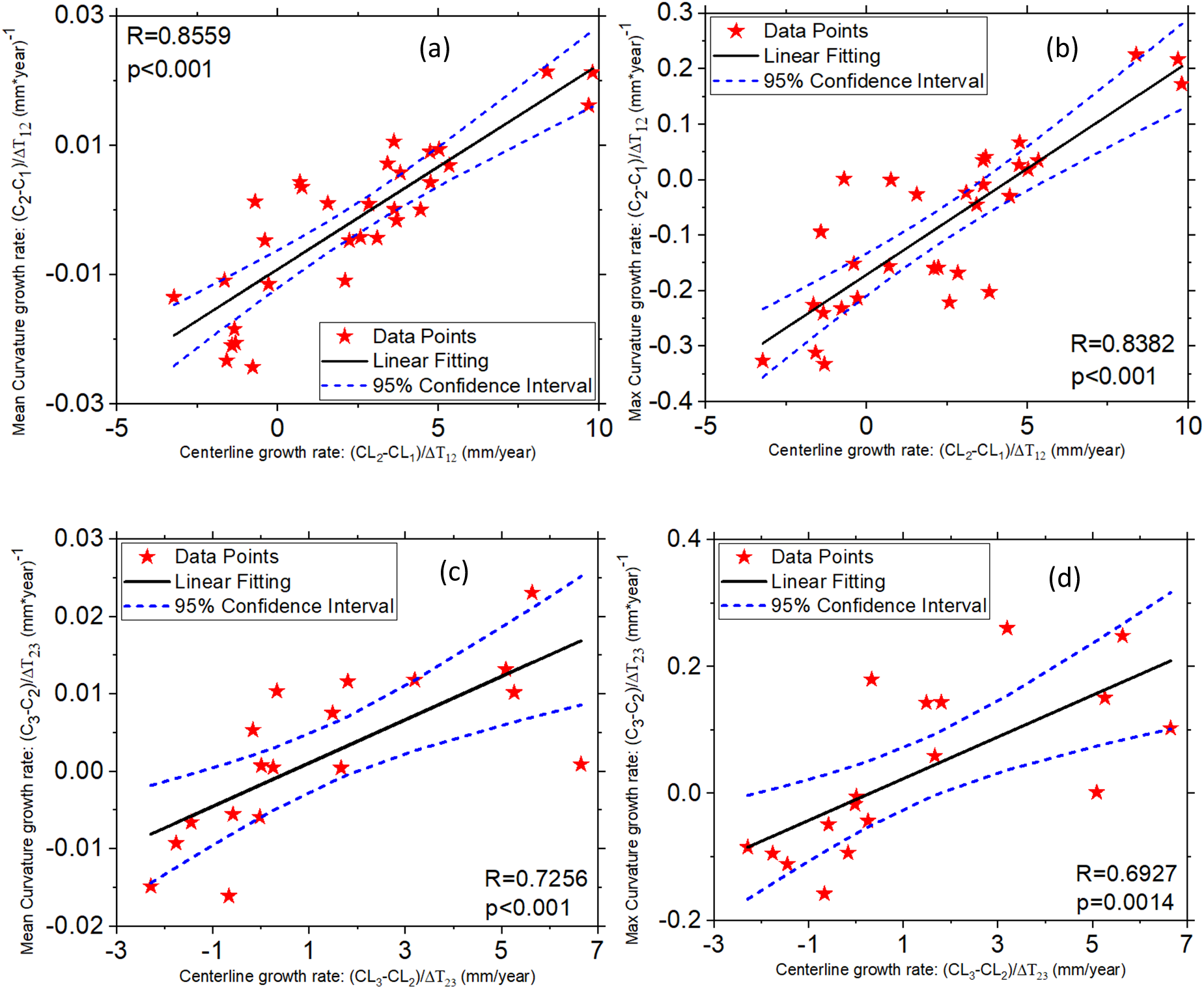
(a, b): Correlations between the centerline length (CL) growth rate and mean/maximum(a/b) centerline curvature (CC) growth rate during Scan-1&Scan-2; (c, d) Correlations between the CL growth rate and mean/maximum CC growth rate during Scan-2&Scan-3.

### 3.5 Diameter growth (CT-1 & CT-2) vs. diameter growth (CT-2 & CT-3)

We also investigated the relationship between diameter growth rates calculated from CT-1 & CT-2, and from CT-2 & CT-3. The results (Fig. 5) showed that they have positive correlations for both mean (*R* = 0.6004, *p* = 0.0084) and maximum (*R* = 0.4158, *p* = 0.0862) values. By comparing these results with those in Figs. 2e, f and Figs. 3e, f, we found that early centerline growth (CT-1 & CT-2) predicted later diameter growth (CT-2 & CT-3) more strongly (|*R*| > 0.72) than did early diameter growth itself (|*R*| < 0.61), indicating the centerline may be better predictors for diameter growth of chronic TBAD.

**Fig. 5.**
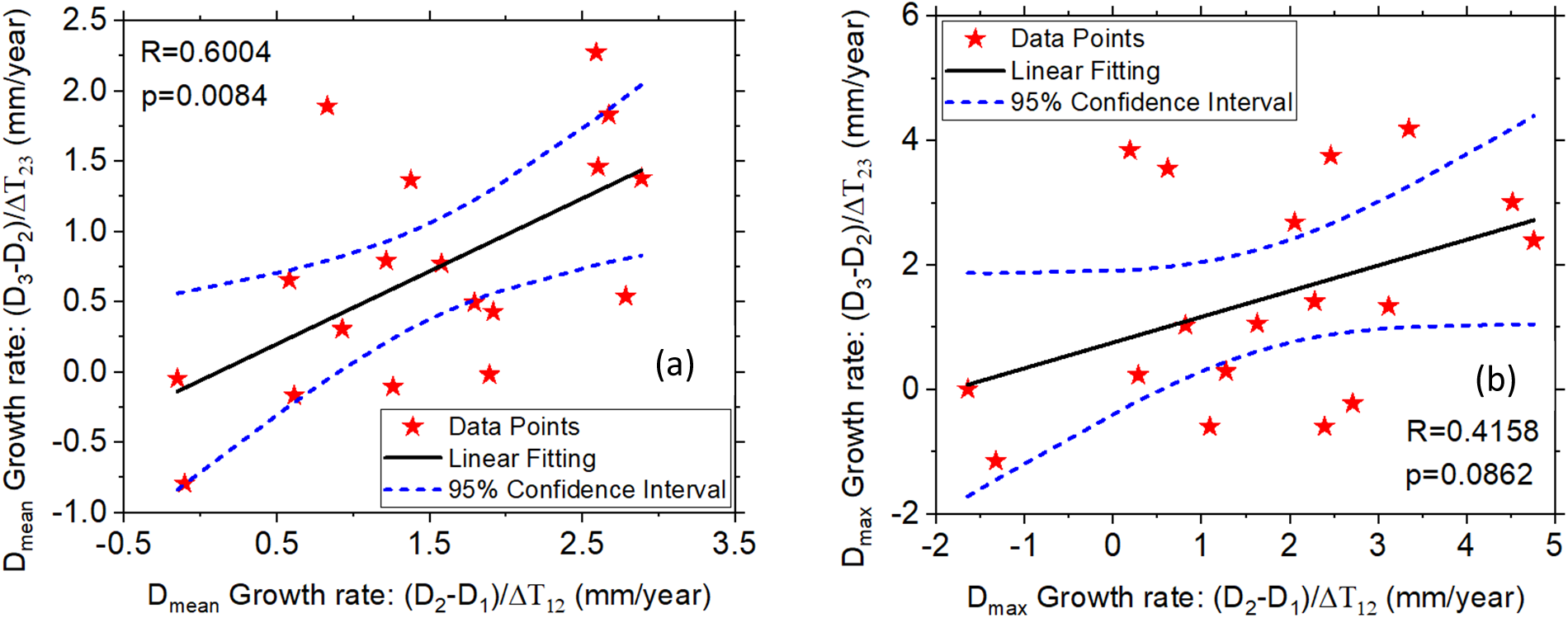
Correlations between the diameter growth rate of Scan-1&Scan-2 and Scan-2&Scan-3: (a) mean and (b) maximum values.

### 3.6 Volume changes

Volumetric analysis of the descending thoracic aorta (left subclavian to celiac) showed substantial expansion across the cohort. Patients were stratified based upon a change in maximal descending thoracic aorta of 5mm (Δ*D*_max_) between their 1^st^ and last available CT Scans. Group-1 (Δ*D*_max_> 5 mm) had a mean percentage volume increase of 50.10% ± 4.89% compared to Group-2 (Δ*D*_max_< 5 mm) which had a mean percentage volume increase of 14.60% ± 2.56% (p < 0.001, Fig. 6)). Notably, measurable volume growth was observed even with minimal diameter change, indicating that longitudinal elongation can drive volumetric expansion despite stable diameters.

**Fig. 6.**
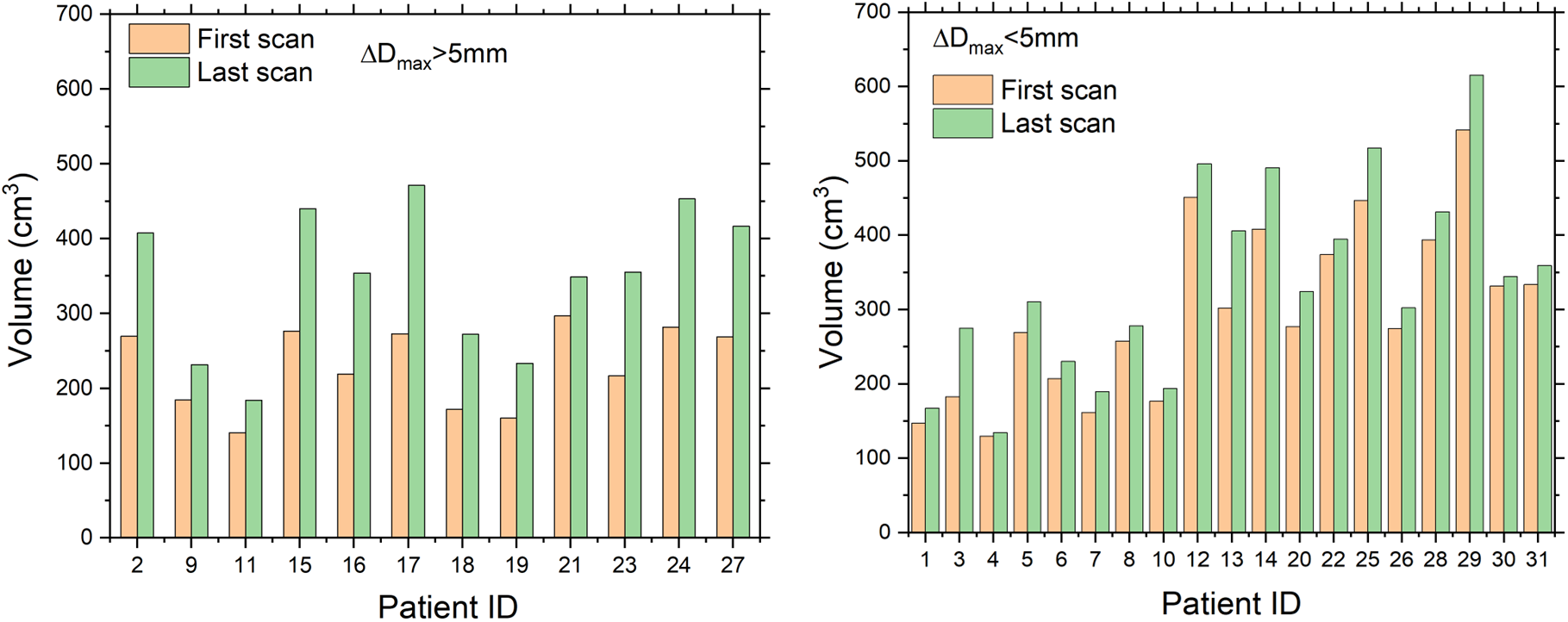
Volume change of the descending thoracic aorta (left subclavian to celiac) between the first and last CT scans. (a) Group-1: maximal diameter change Δ*D*_max_> 5 mm (from the first to the last scan); (b) Group-2: Δ*D*_max_< 5 mm.

## 4. DISCUSSION

A complete understanding of the pathophysiology of TBAD remains elusive. It is unknown why select patients develop lethal false lumen aneurysms requiring surgical intervention in the chronic phase of the disease, while others remain safe with small, stable aortic diameters. Investigations seeking anatomic predictors of aortic growth in TBAD patients have failed to produce any consistent, validated metrics other than aortic diameter^13^. Recent findings that ascending aortic length and curvature may play a role in the prediction of aortic dissection^17^ prompted our group to examine these metrics in patients with uncomplicated TBAD. Using sophisticated three-dimensional imaging analysis techniques, the results of this paper demonstrate that aortic growth in the descending thoracic aorta of TBAD patients occurs not only in the radial direction with diameter expansion, but also in the longitudinal direction with increased length and tortuosity. Furthermore, the growth rate in the longitudinal direction has an inverse relationship to growth in the radial direction.

Using three-dimensional aortic reconstructions of patients with TBAD and customized MATLAB code, we performed 50 separate aortic diameter measurements on the descending thoracic aorta based upon planes orthogonal to the medial axis of the aorta. This axis is also known as the aortic centerline. Aortic centerline measurements are a well-recognized method that allows for accurate aortic diameters to be measured perpendicular to the perceived aortic blood flow channel. The centerlines were also used to assess aortic growth in the longitudinal direction (Centerline Length) and evaluate changes in aortic tortuosity (Centerline Curvature).

Using this detailed aortic centerline analysis, our data demonstrates that the dissected descending thoracic aorta grows in both the radial and longitudinal directions, resulting in diameter expansion and centerline elongation respectively. Elongation of the aortic centerline indicates aortic growth in the longitudinal direction. Since the descending thoracic aorta is mechanically tethered by the ligamentum arteriosum proximally and the aortic hiatus distally, the aorta can bend as it elongates, resulting in increased tortuosity^15^. This hypothesis is supported by the positive correlation observed between centerline length and centerline curvature growth rates (Fig. 4). In this study, we measured tortuosity by calculating the centerline curvature, which increased in 12 patients in this study. Analysis of the relationships between growth in the longitudinal and radial directions revealed an inverse growth relationship between centerline length and aortic diameter. Aortic tortuosity or centerline curvature was also found to have an inverse relationship to aortic diameter.

Interpretation of this data explains why select patients being treated with OMT and surveillance imaging can appear on analysis of their maximum aortic diameter to be stable and mistakenly be categorized as “not growing”. Based upon the data of the current manuscript, the aortic is likely growing in the longitudinal direction and not the radial direction, resulting in a stable aortic diameter, but increased aortic centerline length, and an increased aortic volume. The inverse correlations of growth observed between centerline length and curvature compared to diameter (Figs. 2-3) indicate that the more the aorta grows in the longitudinal direction (i.e., centerline elongation), the less it grows in the circumferential direction (i.e., diameter enlargement). The results (Figs. 2-4) also demonstrated that early centerline growth predicted later diameter growth more strongly than did early diameter growth itself.

An important implication of the findings is the potential for early centerline-derived metrics to enhance individualized surveillance strategies in chronic TBAD. While current clinical guidelines rely heavily on absolute aortic diameter and diameter growth rate to guide intervention^25,26^, this study shows that early changes in centerline length and curvature strongly predict future diameter expansion. The observation suggests that geometric remodeling precedes and potentially drives aneurysmal progression, supporting the utility of centerline length and curvature as early markers of disease progression. Incorporating these metrics into clinical workflows—especially when derived from routine surveillance CT scans—may enable a more proactive and personalized approach to patient management, identifying those at higher risk before critical thresholds are reached.

In a broader biomechanical context, our results underscore the complex three-dimensional nature of aortic growth and remodeling (G&R) in chronic TBAD. It has been shown^27–29^ that aortic wall G&R is strongly influenced by local mechanical stimuli such as aortic wall stress. Both circumferential and longitudinal wall stresses may play distinct roles in modulating tissue G&R, potentially driving diameter expansion and centerline elongation, respectively. Understanding these directional dependencies is essential for interpreting how geometric markers such as curvature and length evolve over time. Future studies may investigate the spatial-temporal relationship between wall stress components and aortic shape change in TBAD, leveraging imaging-based stress analysis and longitudinal follow-up data. Furthermore, advanced computational modeling frameworks^30–32^ may be employed to simulate and predict patient-specific G&R trajectories. Integrating the centerline metrics into such models would provide a mechanistic foundation for evaluating disease progression and optimizing the timing of surgical intervention.

Limitations of the current study include variability in the imaging follow-up and a small sample size. The time intervals between successive CT scans that were available for analysis are highly variable among patients, which introduces temporal inconsistency in the longitudinal analysis. Ideally, uniform scan intervals would allow for more precise comparisons of growth trends. Secondly, the sample size of this study is relatively small (n = 31), which may limit the statistical power of our findings.

## 5. CONCLUSIONS

This study demonstrates that the growth of the aortic centerline length and curvature has an inverse relationship with aortic diameter growth in the chronic phase of Type B Aortic Dissection, and early centerline length and curvature growth predicted later diameter growth more strongly than did early diameter growth itself. The centerline length and curvature may be novel morphologic metrics that can be used to predict diameter growth and applied as early markers of the disease progression in chronic TBAD.

## Data Availability

All data produced in the present work are contained in the manuscript

## Glossary of Abbreviations

TBAD: type B aortic dissection
CT: computed tomography
3D: three-dimensional
TEVAR: thoracic endovascular aortic repair
OMT: optimal medical therapy
CL: centerline length
CC: centerline curvatures
FL: false lumen

## Acknowledgments

This study is supported by NIH R01HL155537 and grants from the Carlyle Fraser Heart Center.

The author H.D. thanks Kristina Porte for her assistance on data collections.

## References

1. MacGillivray TE, Gleason TG, Patel HJ, et al. The Society of Thoracic Surgeons/American Association for Thoracic Surgery clinical practice guidelines on the management of type B aortic dissection. The Journal of Thoracic and Cardiovascular Surgery. 2022;163(4):1231–1249.

2. Tadros RO, Tang GH, Barnes HJ, et al. Optimal treatment of uncomplicated type B aortic dissection: JACC review topic of the week. Journal of the American College of Cardiology. 2019;74(11):1494–1504.

3. Rasiah MG, Abdelhalim MA, Modarai B. Need for and update on clinical trials for uncomplicated type B aortic dissection. JVS-Vascular Insights. 2024;2:100130.

4. Nooromid M, Creisher BA, Abai B. Treatment of uncomplicated type B aortic dissection: optimal medical therapy vs TEVAR+ optimal medical therapy. Vascular and endovascular surgery. 2024;58(1):115–122.

5. Xiang D, Kan X, Liang H, et al. Comparison of mid-term outcomes of endovascular repair and medical management in patients with acute uncomplicated type B aortic dissection. The Journal of thoracic and cardiovascular surgery. 2021;162(1):26–36. e1.

6. Nakamura K, Kobayashi K, Nakai S, et al. Safe and favorable prognosis of thoracic endovascular aortic repair for the low-risk patients with non-acute type B aortic dissection. Frontiers in Cardiovascular Medicine. 2024;11:1442800.

7. Luebke T, Brunkwall J. Type B Aortic Dissection: A Review of Prognostic Factors and Meta-analysis of Treatment Options. Aorta, 2, 265–278. 2014.

8. Ali A, Khwaja S, Saavedra J, et al. Experience with Thoracic EndoVascular Aortic Repair (TEVAR) treatment of uncomplicated Stanford type B aortic dissection, 2022 Updates. Medical Research Archives. 2022;10(12)

9. van Bakel TM, Arthurs CJ, Nauta FJ, et al. Cardiac remodelling following thoracic endovascular aortic repair for descending aortic aneurysms. European Journal of Cardio-Thoracic Surgery. 2019;55(6):1061–1070.

10. Mandigers TJ, Bissacco D, Domanin M, et al. Cardiac and aortic modifications after endovascular repair for blunt thoracic aortic injury: a systematic review. European Journal of Vascular and Endovascular Surgery. 2022;64(2-3):176–187.

11. Yuan K, Potluri VK, Gorantla A, et al. Cardiac remodeling and antihypertensive medication changes after thoracic endovascular aortic repair vs open surgical repair. Journal of Vascular Surgery. 2025;81(3):566–573.

12. Dong Z, Yang H, Li G, et al. Preoperative predictors of late aortic expansion in acute type B aortic dissection treated with TEVAR. Journal of Clinical Medicine. 2023;12(8):2826.

13. Spinelli D, Benedetto F, Donato R, et al. Current evidence in predictors of aortic growth and events in acute type B aortic dissection. Journal of vascular surgery. 2018;68(6):1925–1935. e8.

14. Romeiro AIB. Predictors of adverse events in uncomplicated type B aortic dissection: a systematic review with meta-analysis. PQDT-Global. 2021;

15. Schwartz SI, Durham C, Clouse WD, et al. Predictors of late aortic intervention in patients with medically treated type B aortic dissection. Journal of vascular surgery. 2018;67(1):78–84.

16. Ante M, Mylonas S, Skrypnik D, et al. Prevalence of the computed tomographic morphological DISSECT predictors in uncomplicated Stanford type B aortic dissection. European Journal of Vascular and Endovascular Surgery. 2018;56(4):525–533.

17. Wu J, Zafar MA, Li Y, et al. Ascending aortic length and risk of aortic adverse events: the neglected dimension. Journal of the American College of Cardiology. 2019;74(15):1883–1894.

18. Sun L, Li X, Wang G, et al. Relationship between length and curvature of ascending aorta and type A dissection. Frontiers in Cardiovascular Medicine. 2022;9:927105.

19. Sun L, Li J, Wang L, et al. Aortic geometric alteration associated with acute type B aortic dissection: angulation, tortuosity, and arch type. Frontiers in Physiology. 2021;12:708651.

20. Eliathamby D, Gutierrez M, Liu A, et al. Ascending aortic length and its association with type A aortic dissection. Journal of the American Heart Association. 2021;10(13):e020140.

21. Geronzi L, Haigron P, Martinez A, et al. Assessment of shape-based features ability to predict the ascending aortic aneurysm growth. Frontiers in Physiology. 2023;14:1125931.

22. Sun L, Li H, Feng X, et al. Morphological risk of acute type A aortic dissection in the mildly to moderately dilated aorta. European Journal of Cardio-Thoracic Surgery. 2024;65(1):ezae016.

23. Fleischmann D, Afifi RO, Casanegra AI, et al. Imaging and surveillance of chronic aortic dissection: a scientific statement from the American Heart Association. Circulation: Cardiovascular Imaging. 2022;15(3):e000075.

24. Do Carmo MP. Differential geometry of curves and surfaces: revised and updated second edition. Courier Dover Publications; 2016.

25. Czerny M, Grabenwoeger M, Berger T, et al. EACTS/STS Guidelines for diagnosing and treating acute and chronic syndromes of the aortic organ. European Journal of Cardio-Thoracic Surgery. 2024;65(2):ezad426.

26. Isselbacher EM, Preventza O, Hamilton Black III J, et al. 2022 ACC/AHA guideline for the diagnosis and management of aortic disease: a report of the American Heart Association/American College of Cardiology Joint Committee on Clinical Practice Guidelines. Journal of the American College of Cardiology. 2022;80(24):e223–e393.

27. Rachev A, Stergiopulos N, Meister J-J. Theoretical study of dynamics of arterial wall remodeling in response to changes in blood pressure. Journal of Biomechanics. 1996;29(5):635–642.

28. Mousavi SJ, Farzaneh S, Avril S. Patient-specific predictions of aneurysm growth and remodeling in the ascending thoracic aorta using the homogenized constrained mixture model. Biomech Model Mechanobiol. 2019;18:1895–1913.

29. Dong H, Liu M, Qin T, et al. Engineering analysis of aortic wall stress and root dilatation in the V-shape surgery for treatment of ascending aortic aneurysms. Interactive cardiovascular and thoracic surgery. 2022;

30. Humphrey J, Rajagopal K. A constrained mixture model for growth and remodeling of soft tissues. Mathematical models and methods in applied sciences. 2002;12(03):407–430.

31. Dong H, Sun W. A novel hyperelastic model for biological tissues with planar distributed fibers and a second kind of Poisson effect. Journal of the Mechanics and Physics of Solids. 2021;151:104377.

32. Dong H, Liu M, Qin T, et al. A novel computational growth framework for biological tissues: Application to growth of aortic root aneurysm repaired by the V-shape surgery. Journal of the Mechanical Behavior of Biomedical Materials. 2022:105081.

